# Timeline of SARS-CoV2 spread in Italy: results from an independent serological retesting

**DOI:** 10.1101/2021.07.14.21260491

**Authors:** Emanuele Montomoli, Giovanni Apolone, Alessandro Manenti, Mattia Boeri, Paola Suatoni, Federica Sabia, Alfonso Marchianò, Valentina Bollati, Ugo Pastorino, Gabriella Sozzi

**Affiliations:** University of Siena, Siena, Italy; VisMederi S.r.l., Siena, Italy; Fondazione IRCCS Istituto Nazionale Tumori, Milan, Italy; VisMederi Research S.r.l., Siena, Italy; Epidemiology, Epigenetics and Toxicology Lab, University of Milan, Milan, Italy

**Keywords:** SARS-CoV-2, antibodies

## Abstract

The massive emergence of COVID19 cases in the first phase of pandemic within an extremely short period of time suggest that an undetected earlier circulation of SARS-CoV-2 might have occurred, as documented by several papers in different countries, including a few that reported positive cases even earlier the first cases identified in Wuhan. Given the importance of this evidence, an independent evaluation was recommended. Here we report the results of SARS-CoV-2 antibodies blind retesting of blood samples collected in the prepandemic period in Italy, and in control samples collected one year before, by two independent centers. Results suggest the presence of SARS-CoV-2 antibodies in some samples collected in the prepandemic period, though the detection of IgM and/or IgG binding and neutralizing antibodies is strongly dependent on the different serological assays and thresholds employed, while being absent in control samples collected one year before. These findings highlight the importance of harmonizing serological assays for testing SARS-CoV-2 virus spreading and may contribute to a better understanding the future virus dynamics.

**Article Summary Line:** We report the results of an independent retesting of SARS-CoV-2 antibodies in blood samples collected in prepandemic period in Italy and in matched samples collected one year before. The findings indicate the presence of IgM and/or IgG antibodies in selected samples of the prepandemic period only with different performance of serological assays used by the two centers. The results could give highlights on SARS-CoV-2 circulation in the pre-pandemic period and contribute to better predict future virus dynamic.

## Introduction

The timeline of the first COVID-19 cases remains an unanswered question (1,2). The first declared case of COVID-19 worldwide was dated on December 8, 2019, in Wuhan, China. In Europe, although local transmission was only identified in the second half of February in most countries, there is accumulated evidence that SARS-CoV-2 circulated in earlier times, even earlier that the first cases identified in Wuhan.

This hypothesis is supported by a bunch of published studies including environmental waste water testing (3,4) as well as seroprevalence and molecular analyses on retrospectically analyzed clinical samples of asymptomatic and symptomatic subjects. Positive SARS-CoV-2 RT-PCR results were reported in France in a patient with pneumonia on December 27^th^, 2019 (5) and in a child with suspected measles and a woman with extended skin dermatosis on November 2019, in Milan, Italy (6,7).

Large seroprevalence studies, in USA and in Europe support an earlier than expected circulation of the virus. Retrospective SARS-CoV-2 serological testing of 7,389 routine blood donations collected in nine U.S. states from December 13, 2019-January 17, 2020 suggested that the virus was present as early as December 13–16, 2019 (8).

Using serum samples routinely collected in 9144 adults from a French general population-based cohort, 353 participants with a positive anti-SARS-CoV-2 IgG test were identified. Notably, 13 participants with positive ELISA-S had been sampled between November 5, 2019 and January 30, 2020 and were confirmed by neutralizing antibodies testing. In these positive subjects, the authors identified symptoms, history of possible exposure, or specific events compatible with early SARS-CoV-2 infection (9).

The Italian Ministry of Health accomplished a large SARS-CoV-2 seroprevalence study in a representative sample of 64,660 subjects collected between May 25^th^ and July 15^th^, 2020. A global prevalence rate of 2.5% was reported, with a peak in the Lombardy region (7.5%) and in particular in Bergamo Province (24%) (www.salute.gov.it). According to these numbers, the true number of Italians who had been in contact with the virus would be approximately 1.5 million, many of which asymptomatic, an estimate almost 5 times higher than the official figures reported suggesting an under surface SARS-CoV-2 circulation.

We investigated the presence of SARS-CoV-2 Receptor-Binding Domain (RBD)-specific antibodies in blood samples of 959 asymptomatic individuals enrolled in the SMILE prospective lung cancer screening trial (clinicaltrials.gov ID: NCT03654105) between September 2019 and March 2020, across all the Italian regions. SARS-CoV-2 IgM/IgG RBD-specific antibodies were detected in 111 of 959 (11.6%) subjects, starting from September 2019 (14%), with a cluster of positive cases (>30%) on the 2nd week of February 2020 and the highest number (53.2%) in Lombardy (10). Twelve subjects with samples collected on July 2019 were also included in the study and resulted all negatives (data not shown). The publication of our report generated a lively debate on the possibility that the virus circulated months earlier in Italy without the surveillance programs were able to identify any signs of its presence. To validate these findings, we were offered to blindly retest our samples in an external WHO affiliated laboratory (Erasmus Medical Center, Rotterdam) by using different serological assays. Here we report the results of this cross-validation study.

## Materials and Methods

The sample series included: 29 plasma samples collected in the SMILE trial (clinicaltrials.gov ID: NCT03654105) starting from July 23, 2019 until February 17, 2020 (10); 29 plasma samples from another lung cancer screening cohort (bioMILD, clinicaltrials.gov ID: NCT02247453) collected from July 14, 2018 until February 23, 2019, matched by date of collection, sex, age, smoking habits. The Institutional Review Board and Ethics Committee of Fondazione IRCCS Istituto Nazionale dei Tumori of Milan approved the study. All eligible subjects provided written informed consent. Nine samples from asymptomatic and symptomatic convalescent COVID19 patients with positive molecular (RT-PCR) swab collected from the National Institute for Biological Standard and Control (NIBSC), the University of Siena and the University of Milan, as well as 2 commercial (BioIVT) negative control samples were also included (Table 2).

**Table 1.** Serological results on the SMILE series 2019-2020

| Date of collection | Italian region | IgG |  | IgM |  | Neutralization assay |  |
| --- | --- | --- | --- | --- | --- | --- | --- |
|  |  | VisMederi | Erasmus | VisMederi | Erasmus | MN VisMederi | PRNT Erasmus |
| 23/07/2019 | N/A | Neg | Neg | Neg | Neg | N/A | N/A |
| 24/07/2019 | N/A | Neg | Neg | Neg | Neg | N/A | N/A |
| 03/09/2019 | Lombardy | Neg | Neg | Neg | Neg | N/A | N/A |
| 03/09/2019 | Veneto | Neg | Neg | Pos (RBD) | Neg | Neg | N/A |
| 05/09/2019 | Liguria | Neg | Neg | Borderline | Neg | Neg | N/A |
| 10/09/2019 | N/A | Neg | Neg | Neg | Neg | N/A | N/A |
| 12/09/2019 | Lombardy | Neg | Neg | Neg | Neg | N/A | N/A |
| 17/09/2019 | N/A | Neg | Neg | Neg | Neg | N/A | N/A |
| 02/10/2019 | Piedmont | Pos (RBD) | Neg | Neg | Neg | Neg | Neg |
| 07/10/2019 | Valle'd'Aosta | Neg | Neg | Pos (RBD) | Neg | Pos | N/A |
| 07/10/2019 | Liguria | Pos (RBD) | Neg | Neg | Neg | Neg | Neg |
| 07/10/2019 | Lombardy | Neg | Neg | Pos (RBD) | Neg | Pos | N/A |
| 08/10/2019 | Lombardy | Neg | Neg | Pos (RBD) | Neg | Pos | N/A |
| 10/10/2019 | Lombardy | Pos (RBD) | Neg | Neg | Neg | Neg | Neg |
| 10/10/2019 | Lombardy | Neg | Neg | Pos (RBD) | Pos (RBD) | Neg | Neg |
| 15/10/2019 | Lombardy | Neg | Neg | Pos (RBD) | Neg | Neg | N/A |
| 15/10/2019 | N/A | Neg | Neg | Neg | Neg | N/A | Neg |
| 21/10/2019 | Tuscany | Neg | Neg | Pos (RBD) | Neg | Pos | N/A |
| 22/10/2019 | Lombardy | Neg | Neg | Neg | Neg | N/A | N/A |
| 07/11/2019 | Lombardy | Neg | Neg | Borderline | Neg | Pos | N/A |
| 11/11/2019 | Lombardy | Neg | Pos (S1) | Pos (RBD) | Borderline | Neg | Neg |
| 26/11/2019 | Lazio | Pos (RBD) | Neg | Neg | Neg | Neg | Neg |
| 13/12/2019 | Lombardy | Pos (RBD) | Neg | Borderline | Neg | Neg | Neg |
| 17/12/2019 | Campania | Pos (RBD) | Pos (S1) | Pos (RBD) | Neg | Neg | Neg |
| 18/12/2019 | Liguria | Neg | Neg | Pos (RBD) | Neg | Neg | N/A |
| 28/01/2020 | Lombardy | Pos (RBD) | Pos (S1,NP) | Neg | Neg | Neg | Neg |
| 05/02/2020 | Lazio | Neg | Neg | Pos (RBD) | Pos (RBD) | Pos | Pos |
| 12/02/2020 | Lombardy | Neg | Neg | Pos (RBD) | Neg | Neg | Neg |
| 17/02/2020 | Piedmont | Neg | Neg | Pos (RBD) | Neg | Neg | N/A |

RBD: Receptor-Binding Domain; NP: Nucleocapsin Protein.

**Table 2.** Serological results in negative controls, convalescent symptomatic and asymptomatic COVID19 patients

| Sample | Center of collection | IgG |  | IgM |  | Neutralization |  |
| --- | --- | --- | --- | --- | --- | --- | --- |
|  |  | VisMederi | Erasmus | VisMederi | Erasmus | MN VisMederi | PRNT Erasmus |
| Negative control | NIBSC | Neg | Neg | Neg | Neg | Neg | N/A |
| Negative control | NIBSC | Neg | Neg | Neg | Neg | Neg | N/A |
| Convalescent symptomatic | NIBSC | Pos (RBD) | Pos (S1,ecto,NP) | Neg | N/A | Pos | Pos |
| Convalescent symptomatic | BioIVT | Pos (RBD) | Pos (S1,ecto,NP) | Pos (RBD) | Pos (RBD) | Pos | Pos |
| Convalescent symptomatic | BioIVT | Pos (RBD) | Pos (S1,ecto,NP) | Pos (RBD) | Neg | Pos | Pos |
| Convalescent asymptomatic | UNISI | Pos (RBD) | Neg | Neg | Neg | Neg | N/A |
| Convalescent asymptomatic | UNIMI | Pos (RBD) | Neg | Neg | N/A | Neg | Neg |
| Convalescent asymptomatic | UNIMI | Pos (RBD) | Neg | Neg | Neg | Neg | N/A |
| Convalescent asymptomatic | UNIMI | Pos (RBD) | Neg | Neg | Neg | Neg | N/A |
| Convalescent asymptomatic | UNIMI | Pos (RBD) | Neg | Neg | Neg | Neg | N/A |
| Convalescent asymptomatic | UNIMI | Pos (RBD) | Neg | Neg | N/A | Neg | Neg |

NIBSC: National Institute for Biological Standard and Control; UNISI: University of Siena; UNIMI: University of Milan; RBD: Receptor-Binding Domain; NP: Nucleocapsin Protein.

VisMederi, an independent laboratory part of the CEPI consortium (https://cepi.net), used proprietary IgG-RBD/IgM-RBD Elisa assays and a qualitative Micro-Neutralization CPE-Based assay (MN) with the aim to increase the sensitivity of the test (11,12). The assays were qualified and validated following the guideline issued by the International Council for Harmonization of Technical Requirements for Pharmaceuticals for Human Use (ICH) (www.ich.org). A cut-off value of each plate was obtained by multiplying 3 times the “BLANK” optical density (OD) signal derived by six micro-wells containing sample diluents and secondary HRP-antibody, but no analyte (12). By Vismederi standards, criteria of RBD-specific IgM and/or IgG above the cut-off value, with or without the presence of neutralizing antibodies, indicate presence of SARS-CoV-2 specific antibodies (12).

Erasmus tested the samples with a multiplex IgG protein array including three antigens: S1, ecto and nucleocapsid protein (NP). By Erasmus standards, criteria of IgG triple antigen positives, with confirmation of neutralization assay using Plaque Reduction Neutralization test (PRNT), were needed before scoring a sample as positive. For IgM detection, Erasmus adopted the Elisa IgM commercial kit Wantai using cut-off as suggested by the manufacturer (13,14).

Specificity of both VisMederi and Erasmus assays was validated across the most common HCoV sera (12,13).

For both the VisMederi and Erasmus assays, the IgM results were calculated by relating each specimen OD value to the respective cut-off value of the plate. Results were thus expressed as OD ratio and considered positive when > 1. For correlation analysis of nonparametric data, the Spearman rho coeficent (r) with respective 95% confidence interval and two-tailed p-value was calculated. To compare data distributions, p-values were calculated using the Mann-Whitney test. For the analysis and the graphic design the GraphPad Prism software (version 5.2) was adopted.

## Results

According to Vismederi of the 29 SMILE samples, 7 cases were positive for IgG, 16 cases were classified as positive (n=13) or borderline (n=3) for IgM, of which 6 samples were also positive in qualitative microneutralization assay and 6 cases were negative (Table 1). According to the Erasmus Medical Center none of the SMILE series showed IgG triple antigen positivity (3 samples had S1 or S1+NP positivity only), 2 samples were positive and one was in grey zone for IgM detection using Wantai cut-offs (Table 1). Out of the 12 tested samples for the PRNT assay, 1 IgM positive sample has also neutralizing antibodies (Table 1). In 9 additional samples, classified as IgM positives by VisMederi, a signal was also detected by Erasmus, but below the threshold level of the Wantai test (*Figure 1A*). These differences, especially in IgM outcomes, would seem to derive from a different setting of the cut-off values used to attribute positivity or negativity to anti SARS-CoV-2 antibodies. In fact, the ELISA cut-off of VisMederi was established internally through a blind study of symptomatic and asymptomatic subjects positive to molecular swab (15). On the other hand, the Wantai ELISA commercial Kit uses a different cut-off set up for diagnostic purposes in symptomatic patients. Despite this, a significant correlation among IgM values (VisMederi vs. Erasmus) was observed (Spearman r=0.6130; p=0.0004), when OD ratios were considered as continuous variables (*Figure 1A*).

**Figure 1.**
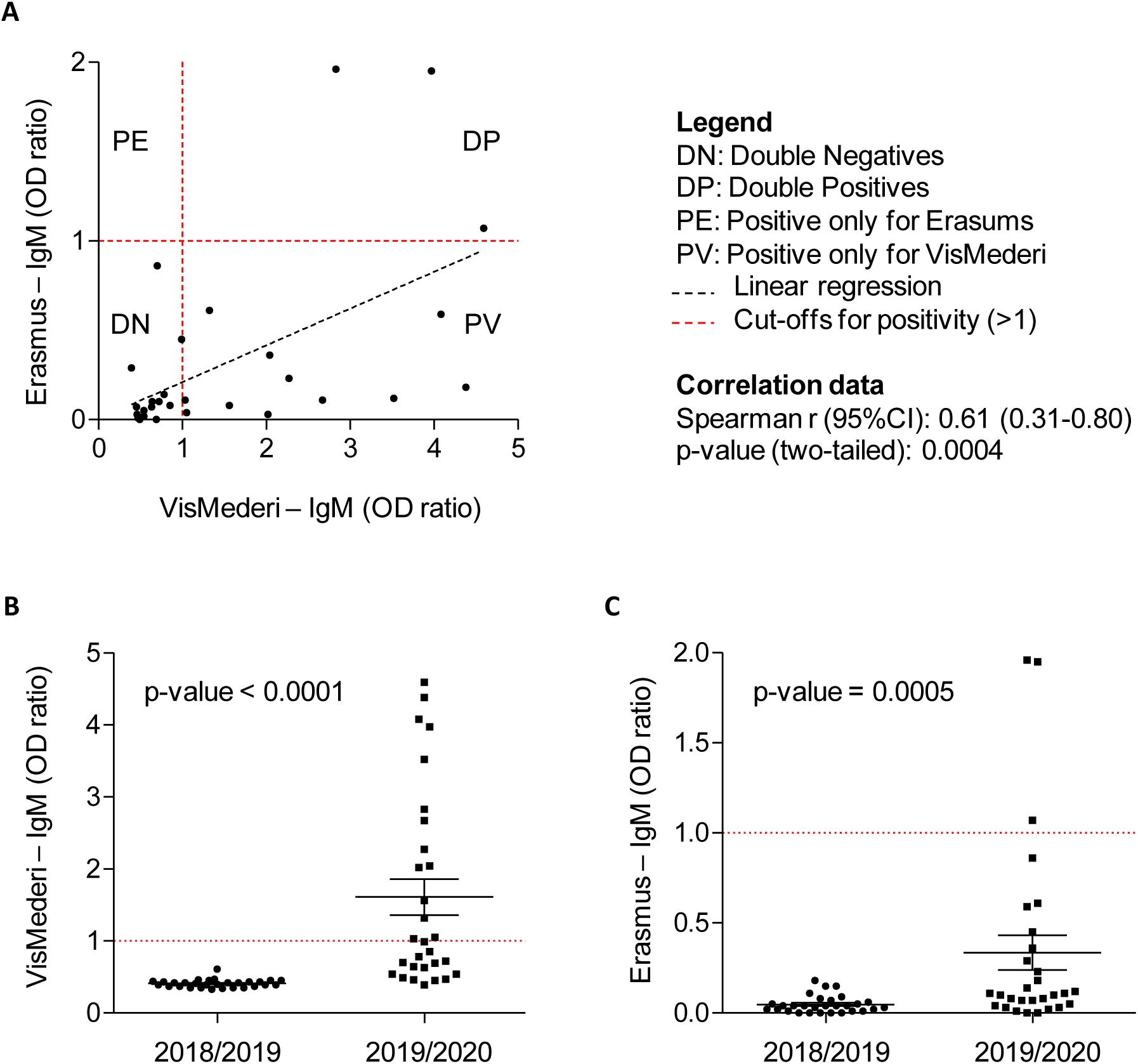
Comparison of IgM data between VisMederi and Erasmus. **(A)** The scatter plot shows the correlation between IgM values (OD ratios) obtained in the two laboratories, considering the 29 selected samples collected from asymptomatic subjects enrolled in the SMILE lung cancer screening trial between July 2019 and February 2020. The dot plots show the distribution of IgM values (OD ratio) obtained by **(B)** VisMederi and **(C)** Erasmus in 29 samples of the SMILE screening trial enrolled in 2019/2020 and 29 matched lung cancer screening volunteers enrolled in the same months of 2018/2019. Horizontal black bars indicate mean value ± standard error. Mann-Whitney p-values are reported.

To better assess assays specificity, 29 plasma samples collected from July 2018 until February 2019 from subjects matched to those of the SMILE study and supposed to be negative were also included. Indeed, all these samples resulted negatives by both VisMederi and Erasmus IgG and IgM assays. By plotting IgM values recorded by both centers a significantly higher distribution of values was observed in samples collected in 2019-2020 versus control samples collected one year before in 2018-2019: p<0.0001 and p=0.0005 by VisMederi and Erasmus, respectively (*Figure 1B-C*).

To have better insights about the assays performances and the concordance between the two laboratories, 11 among positives convalescent patients (3 symptomatic and 6 asymptomatic) and negatives controls (n=2) were included in the study (Table 2). RBD specific IgG were detected by VisMederi in all 9 positive symptomatic and asymptomatic COVID19 subjects whereas only the 3 symptomatic convalescents samples were classified as triple positive in multiple IgG protein array by Erasmus. None of the analyzed samples from asymptomatic convalescents tested positive in neutralization assays whereas the 3 samples from symptomatic convalescents showed neutralizing activity by both Vismederi and Erasmus (titers: 1280, 40, 40 respectively) laboratories.

## Discussion

Overall, the results of this blind retesting indicate the presence of SARS-CoV-2 antibodies in some SMILE samples collected in the prepandemic period. The oldest samples found positive for IgM by both laboratories were collected on October 10^th^, 2019 (Lombardy), November 11^th^, 2019 (Lombardy) and February 5^th^, 2020 (Lazio), the latter with neutralizing antibodies. Two additional samples collected on December 17^th^, 2019 (Campania) and January 28^th^, 2020 (Lombardy) were IgG positive for VisMederi and IgG S1 and IgG S1+NP, respectively, by Erasmus. Additional IgM positive cases could have been detected also by Erasmus by lowering the cut-off of the commercial IgM assay. The older among these putative additional IgM positive samples was collected on September 3^th^, 2019 in the Veneto region, one of the first and mostly affected COVID19 regions.

Noteworthy, particularly for samples from asymptomatic SARS-CoV-2 infected subjects, where the antibody concentration is known to be very low and are rarely able to develop neutralizing antibodies (16–18), the agreement among the different tests used in the two laboratories is poor, highlighting the issue of sensitivity of commercial serological tests. Conversely, samples from symptomatic COVID19 convalescents tested positive for IgG and showed neutralizing activity by both the laboratories. If it is true that high specificity is an important parameter in SARS-CoV-2 seroprevalence studies, even sensitivity is an equally important parameter that should be considered when testing asymptomatic infected subjects.

By Erasmus standards, criteria of IgG triple antigen positives, with confirmation of neutralization assay, before scoring a sample as positive are used because individual antigens do occasionally have some low level reactivity. This is based on in house validation data for surveys of pre-outbreak sera to set a high specificity value for studies in which it is important to be able to have conclusive results. Indeed the percentage of triple positive sera increases starting at 7 days post infection indicating lower sensitivity in recently (<7 days) infected subjects. Accordingly, by using this approach only few symptomatic COVID19 patients were identified while asymptomatic patients and SMILE subjects did not reached the triple antigens cut-off. On the other hand, IgM antibodies and neutralizing activity were found in a SMILE sample collected on February 5^th^, two weeks before the first officially declared COVID19 Italian patient.

Lombardy was the first region to have been affected by the COVID-19 outbreak, with a death rate almost six times higher than in the rest of Italy. A recent large phylogenetic study on 346 SARS-CoV-2 patients in Lombardy allowed the identification of seven SARS-CoV-2 lineages and the presence of local transmission clusters within three of them, suggesting that the virus was circulating undetected for some time before first detection and confirming a central role of Lombardy in the SARS-CoV-2 epidemic (19). Similar conclusions were reached in a previous report in a smaller number of sequenced samples (20). In contrast, no evidence of SARS-CoV-2 viral RNA was found in 1581 respiratory samples collected in the framework of influenza surveillance between October 2019-February 2020 in Lombardy (21). Nonetheless, the presence of neutralizing antibodies in 5 blood donors collected at the beginning of February in the Lodi Red Zone, where the outbreak started, was reported by the same authors (22).

Beyond serological studies and computational genomic phylogenetic analyses the isolation of the SARS-CoV-2 viral genome itself in a respiratory swab of a symptomatic child recovered for suspected measles and in the skin biopsy of a symptomatic woman whit positive IgG Sars-CoV-2 antibodies later on, both in November 2019 in Milan strongly support an early, under surface circulation of the virus (6,7). These findings do not at all suggest that the virus originated in Italy, but endorse the idea that the virus was likely spreading in China before the first known cases and that could be brought around by travelers given the direct connection of China with European and US countries, particularly with the Northern West and East Italian regions, which are among the most industrialized and connected areas of Italy. Support of this hypothesis comes from a comparative genomic analysis of more than 175,000 genomes which delineated 22 distinct SARS-CoV-2 haplogroups with a broad geographic distribution within China pointing towards an early emergence and widespread cryptic circulation of the virus well before its isolation in January 2020 (23). Recently, *Kumar et al*. reconstructed the mutational history of SARS-CoV-2 using a so called ‘mutation order approach’ (MOA) (24). From their analysis of more than 174,000 genomes, major mutational fingerprints revealed useful to identify and track the novel coronavirus spatiotemporal evolution. The progenitor genome identified differed from that of the first coronaviruses sampled in China by three variants, implying that none of the earliest patients represent the index case or gave rise to all the human infections. However, multiple coronavirus infections in China, USA and Europe harbored the progenitor genetic fingerprint in January 2020 and later, suggesting that the progenitor was spreading worldwide months before.

We acknowledge some limitations in this re-test study related to the limited sample size, the highly specific selection of screening participants (heavy smokers ≥ 30 pack years and ≥55 years old), possibly not representative of the general population, and the intrinsic experimental variability of the immunoenzymatic assays employed in the different laboratories. Moreover, we cannot exclude that other confounding conditions, such as preexisting immunity against other agents, might have contributed to the SARS-CoV-2 positivity in our assays. Nonetheless, cross reactivity towards the most common HCoVs was ruled out.

As pointed out in the recent WHO report (www.who.int) and in other commentaries (1,2), studies from different countries suggest that SARS-CoV-2 was growing under the surface for some time before the first diagnosed case in Wuhan. Suggestive evidence from our previous published study (10) and conflicting results from the current retesting exercise, despite adding important signals in this direction, do not allow us to accept or discard this hypothesis. Indeed findings of these studies are only partially confirmed due to the heterogeneity of methods utilized and the risk of non-specific signals in serological assays. Despite that, the report underlines the importance of investigating these potential early events, to solve the still unanswered questions about the origin and timing of the current pandemic and may contribute to a better understanding of the future virus circulation dynamics.

## Data Availability

All data are available upon request to the corresponding author.

## Acknowledgments

The authors thank the Department of Viroscience, Erasmus MC for performing SARS-CoV-2 serology on the shared samples, for providing results and for critical discussion of data. The authors would like to acknowledge the collaboration and support of WHO for the exploitation of this study. The opinions expressed in this article do not necessarily reflect those of Erasmus and WHO.

## Author Bio

**Emanuele Montomoli** is Full Professor on Public Health at University of Siena in Italy, he’s also Founder and Chief Scientific Officer at VisMederi srl in Italy. Primary research interest are: public health, prevention of infectious diseases, vaccines development and vaccines clinical trials. The main expertise is in the area of laboratory assays standardization and validation.

**Giovanni Apolone**, Medical Doctor with post-doctoral degrees in Internal Medicine and Pharmacology, is currently the Scientific Director of the Foundation IRCCS Istituto Nazionale dei Tumori of Milan. His main fields of interest are: Methodological, ethical and regulatory aspects of clinical research, with special emphasis on oncology and anticancer drug, and health care evaluation.

